# Self-Perceived Decline in Memory and Concentration 9 and 12 months post COVID-19 infection

**DOI:** 10.1101/2024.12.19.24319396

**Authors:** Carolina Ruivinho, Marta Moniz, Ana Rita Goes, Patrícia Soares, Andreia Leite, LOCUS group

**Affiliations:** NOVA National School of Public Health, Public Health Research Centre, Comprehensive Health Research Center, CHRC, LA-REAL, CCAL, NOVA University Lisbon, Lisbon, Portugal; Centre for Vectors and Infectious Diseases Research, National Institute of Health Doutor Ricardo Jorge, Águas de Moura, Portugal; Department of Epidemiology, National Institute of Health Doutor Ricardo Jorge, Lisbon, Portugal

**Keywords:** Cognitive decline, Concentration issues, COVID-19 infection, Long Covid, Post-COVID condition, SARS-CoV-2, Memory loss

## Abstract

**Introduction:** COVID-19 infection caused by SARS-CoV-2 has led to significant long-term health challenges, including Long COVID or Post-COVID condition, that can include symptoms such as cognitive decline, memory loss, and concentration issues. This study investigates the prevalence and risk factors of post-COVID cognitive symptoms among individuals tested for COVID-19.

**Methods:** A cross-sectional study was conducted in Lisbon and Tagus Valley, targeting individuals tested for COVID-19 in August 2022. Participants were selected from a random sample of 10,000 individuals. Data were collected via computer-assisted telephone interviews at 9 and 12 months post-test, covering sociodemographic details, health behaviors, pre-existing conditions, and COVID-19 symptoms. The primary outcome was the presence of at least one cognitive symptom (memory loss and/or concentration issues) at 9 and 12 months. Additionally, each symptom was assessed individually, along with a composite outcome of both symptoms concurrently.

**Results:** At 9 months, memory loss was reported by 24.87% of COVID-19 positive cases versus 10.20% of negatives, and concentration issues by 15.45% of positives versus 7.45% of negatives. At 12 months, memory loss prevalence was 16.67% for positives and 9.45% for negatives, while concentration issues were 9.82% for positives and 2.99% for negatives. Additionally, the prevalence of at least one cognitive symptom was 28.24% in positive cases at 9 months compared to 12.16% in negatives, and 17.81% versus 9.95% at 12 months. Female sex was significantly associated with a higher prevalence of cognitive symptoms at both time points.

**Discussion:** These findings underscore the enduring cognitive impact of COVID-19, with significant disparities in cognitive symptoms between COVID-19 positive and negative individuals observed at both 9 and 12 months post-infection. The higher prevalence of memory loss and concentration issues among COVID-19 positives suggests potential neurological sequelae linked to SARS-CoV-2 infection. Notably, the association of female sex with increased cognitive symptom prevalence warrants further investigation into gender-specific vulnerabilities or biological mechanisms underlying these disparities. Addressing these persistent cognitive symptoms is crucial for long-term patient management and underscores the need for targeted interventions and comprehensive post-COVID care strategies to mitigate long-lasting health implications.

## 1 Introduction

The coronavirus disease 2019 (COVID-19), caused by the severe acute respiratory syndrome coronavirus 2 (SARS-CoV-2), has posed significant challenges globally, extending beyond acute illness to encompass a spectrum of long-term sequelae(Soriano et al., 2022). While initially recognized as primarily a respiratory illness, emerging evidence underscores its multiorgan impact, including notable neurological and cognitive manifestations both during and after acute infection(Gonzalez-Fernandez & Huang, 1910; Möller et al., 2023; Soriano et al., 2022). After recovering from COVID-19, some individuals experience persistent symptoms, a condition commonly referred to as post-COVID condition (PCC) or Long COVID. These symptoms, which may appear either newly or persist from the initial illness, typically last for at least two months, cannot be explained by another diagnosis, and include fatigue, breathing difficulties, and cognitive issues(Soriano et al., 2022). Prevalence has been estimated to be between 10% to 70%(Al-Aly et al., 2022; Crook et al., 2021; Davis et al., 2023; Fernández-de-Las-Peñas et al., 2022; Pérez-González et al., 2022; Sahanic et al., 2023; Wu et al., 2021; Yaksi et al., 2022) of previously infected patients, depending on vaccination status, severity of initial disease and several other factors. The impact of COVID-19 extends beyond physical health, significantly affecting various sectors such as psychiatric hospitals and elder care institutions, highlighting the need to address the unique challenges faced by underserved populations during the pandemic. A recent study reported that, out of 19,573 patients hospitalized in psychiatric hospitals across 17 studies from different regions of the world, a pooled mean of 11.9% were diagnosed with COVID-19(Maximiano-Barreto et al., 2024). Additionally, another review that focused on 48 articles related to elder care institutions identified new risk factors that can inform healthcare services aimed at protecting vulnerable residents in nursing homes(Yin et al., 2024).

In the realm of Long COVID research, cognitive symptoms, such as memory loss, concentration issues, and brain fog, have gained prominence. A recent study comparing cognitive abilities across COVID-19 patient groups found that those with persistent symptoms displayed deficits in working and prospective memory tasks, such as remembering appointments and object recall. Additionally, individuals with persistent COVID-19 symptoms reported more frequent everyday memory lapses compared to controls (Espinar-Herranz et al., 2023). Among an array of studies, memory loss emerges as a prevalent cognitive manifestation (11% to 34.5%) (Ahmed et al., 2022; Fernández-de-las-Peñas et al., 2023; Garrigues et al., 2020; Keijsers et al., 2022; Merza et al., 2023; Pilotto et al., 2021; Søraas et al., 2021), while concentration issues exhibit a lower prevalence (2.6% to 31%) and have received comparatively less research attention(Fernández-de-las-Peñas et al., 2023; Keijsers et al., 2022; Søraas et al., 2021). Moreover, hospitalized patients show a higher prevalence of cognitive post-COVID complications, aligning with trends in other symptom categories(Garrigues et al., 2020; Keijsers et al., 2022; Pérez-González et al., 2022). Regarding their trajectory, most studies indicate that these cognitive symptoms decline over time following infection(Baseler et al., 2022; Fernández-de-las-Peñas et al., 2023). Nonetheless, some studies suggest that these symptoms may worsen over time or even emerge only after 12 months post-infection(Fernández-de-las-Peñas et al., 2023; Ling et al., 2024).

The neurotropic potential of COVID-19, along with associated structural brain alterations, respiratory complications, and critical care interventions, can explain memory loss and cognitive issues(Douaud et al., 2022; Taquet et al., 2023). This underscores the imperative for comprehensive approaches in addressing post-infection cognitive sequelae. Despite the growing recognition of neuropsychological symptoms in post-COVID patients, the predominant focus in research remains on physical manifestations, particularly respiratory symptoms. While some studies are emerging to investigate the prevalence of Long COVID cognitive complications, very few delve into their underlying risk factors. Moreover, most existing knowledge focuses on hospitalized or mixed populations and lacks a comparison group. From a public health perspective, representative samples of COVID-19 cases and non-infected comparators allow to estimate the burden of these complications.

This study aims to address existing knowledge gaps by comparing the prevalence of memory and concentration loss between individuals who tested positive and negative for COVID-19 at 9 and 12 months post-test. Additionally, we aim to investigate the factors influencing memory loss and concentration issues at these time-points. Based on the existing literature, we hypothesize that individuals who tested positive for COVID-19 will report higher prevalence rates of memory loss and concentration issues compared to those who tested negative, and that factors such as age, sex, previous infections, and education levels may significantly influence these cognitive outcomes.

## 2 Materials and methods

### 2.1 Study design and data collection

This cross-sectional study targeted individuals residing in Lisbon and Tagus Valley, encompassing one-third of the country’s population spread across urban and rural areas. Residents with positive and negative SARS-CoV-2 test notifications from the National System of Epidemiological Surveillance (SINAVE) in August 2022 were invited to participate. The General Directorate of Health (DGS) provided the research team with participant information in two phases. On February 28, 2023, the data owner initially provided contact information, including names and cellphone numbers, for a randomly selected sample of 10,000 individuals who had undergone SARS-CoV-2 testing. From March 15, 2023, to June 14, 2023, trained interviewers obtained verbal informed consent via telephone, providing participants the opportunity to either accept or decline participation. Upon obtaining consent, we received eligible individuals’ birthdates and test results and applied the questionnaire through a 30- minute computer-assisted telephone interview. Data collection occurred at two time-points: the first approximately nine months after the SARS-CoV-2 test (between June 12 and August 8 of 2023) and the second approximately twelve months after the SARS-CoV-2 test (between September 27 and November 10 of 2023). Calls were scheduled for the most convenient times for the participants, with a maximum of five call attempts at different hours. The questionnaire gathered sociodemographic data, previous comorbidities, COVID-19 care levels (home-based, primary care, emergency department, hospital admission, hospital admission in intensive care), lifestyle behaviors (e.g., alcohol intake, smoking, physical exercise), and symptoms reported during testing and within the seven days before each interview. Individuals were free not to answer any question, and in questions related to symptoms and health conditions, there was also the option “I do not know.

### 2.2 Study population and sample size

We included individuals who underwent an SARS-CoV-2 test in August 2022, resided in Lisbon and Tagus Valley region during the study period, were 18 years old or older, and consented to participate, regardless of their nationality or immigration status. We excluded individuals who: i) did not have a valid landline or mobile phone number registered; ii) were institutionalized (e.g. residential structures for the elderly or prisons); iii) died between the date of the test and the call; iv) had language barriers (languages not covered by the group of translators that were part of the team of investigators) or deafness, as well as advanced states of mental illness or dementia; v) were Portuguese tourists or emigrants on holidays in Portugal; and vi) tested positive for SARS-CoV-2 after August 2022 but before completing the questionnaire, ensuring an equal time between the test and Long COVID assessment for all participants. Further details on the data collection process and study population can be found in the published study protocol(Dinis Teixeira et al., 2023).

### 2.3 Variables

The primary outcome of this study was the presence of at least one cognitive symptom (memory loss and/or concentration issues) assessed at two distinct time-points: 9 and 12 months following the SARS-CoV-2 test. Regarding the selection of the 9- and 12-month time points, these were based on previous studies indicating that long-term cognitive symptoms can persist for a year or longer following COVID-19 infection(Fernández-de-las-Peñas et al., 2023). Our goal was to describe potential variations in the trajectory of cognitive symptoms, specifically memory and concentration problems, over time. By assessing these symptoms at both 9 and 12 months post-infection, we aimed to identify any changes in symptom prevalence or severity as recovery progressed beyond the acute phase of the illness. Additionally, we assessed each symptom individually (memory loss and concentration issues) as well as a composite outcome indicating the presence of both memory loss and concentration issues concurrently at these specified time-points. Symptom data were self-reported using a Yes/No/Unknown format in a questionnaire based on the International Severe Acute Respiratory and Emerging Infection Consortium (ISARIC) and WHO COVID-19 Clinical Characterisation Protocol(Sigfrid et al., 2021). Participants were asked about ’difficulty remembering’ and ’confusion or lack of concentration’ experienced in the seven days prior to the interview. The 7-day recall period was chosen to minimize recall bias and ensure that participants were reporting recent symptoms rather than symptoms experienced in the distant past. Further details can be found in the supplementary materials (S1B) of the published protocol(Dinis Teixeira et al., 2023). We further used information on demographic characteristics: sex (male/female), age (in years), education level (medium, primary education or lower education; secondary education; higher education), worker (yes/no); behavioral and clinical characteristics: alcohol consumption (never/ 2 to 4 times a month or less/ twice a week or more), physical exercise (defined as ≥ 30 minutes daily - yes/no), and pre-existing health conditions (previous COVID-19 infection, previous psychiatric condition); and COVID-19 related factors: number of symptoms at COVID-19 test, level of care needed (hospitalized/non-hospitalized), number of COVID-19 vaccine doses administered at the time of the first interview. Working status was assessed at each time-point, while the remaining data was only collected during the initial interview at 9 months.

### 2.4 Statistical analysis

Categorical data was summarised as frequencies and percentages, and continuous data were presented as mean (minimum and maximum) and median, along with the corresponding interquartile range (IQR) shown as the 25th and 75th percentiles. Prevalence estimates for SARS-CoV-2 positive and SARS-CoV-2 negative participants were calculated for all four outcomes (memory loss, concentration issues, at least one cognitive symptom and both cognitive symptoms concurrently) by dividing the number of individuals reporting specific symptoms (such as concentration issues and memory loss) at 9 months and 12 months post-testing by the total number of participants in each group, accompanied by a 95% confidence interval (95%CI). A sensitivity analysis was performed for all four outcomes, excluding participants over 60 years old due to the greater age-related cognitive decline in this group(Hedden & Gabrieli, 2004). This analysis aimed to assess the variation in the prevalence of post-COVID cognitive symptoms, eliminating the influence of age-related cognitive decline. We further investigated the association between SARS-CoV-2 positivity and cognitive symptoms by calculating the difference in proportions between positive and negative test groups at 9 and 12 months follow-up to investigate the proportion of symptoms explained by Long COVID.

To explore the association between individuals’ characteristics and the primary outcome (presence of at least one cognitive symptom), we considered only the individuals with a positive SARS-CoV-2 test. Sex, age, education level (as a proxy for income), alcohol intake, physical exercise, COVID-19 symptoms status, pre-existing health conditions (COVID-19, psychiatric condition), and the number of COVID-19 vaccine doses were included as independent variables. These variables were selected in accordance with the possible factors identified in the literature that could affect the onset of post-COVID-19 cognitive symptoms(Ahmed et al., 2022; Al-Aly et al., 2021; Crook et al., 2021; Greißel et al., 2024; Hüfner et al., 2022; Merza et al., 2023; Pilotto et al., 2021; Wang et al., 2021). Given that odds ratios can be overestimated in the presence of frequent events, a robust Poisson regression was employed. This method yields more reliable estimations than logistic regression when analyzing binary outcomes from cross-sectional studies(Barros & Hirakata, 2003). Crude and adjusted prevalence ratios (PR) were estimated, alongside respective 95%CI, adjusting for all the aforementioned variables. Overall, missing values were implicitly handled by subsetting the data based on complete cases for each variable included in the analysis. Data were analyzed using R version 4.3.2.

## 3 Results

From the individuals tested in August 2022, we had access to a random sample of 10,000. Due to privacy concerns and our 9-month deadline, we contacted 6,642 people for the initial questionnaire, with 1,229 consenting to participate. Among them, 120 dropped out, 226 were unreachable, and 65 were ineligible (due to COVID-19 reinfection between the date of the test and the questionnaire date). Thus, a final sample of 818 participants was included in the 1st follow-up analysis. Within this group, 563 tested positive for SARS-CoV-2. After completing the first survey, 47 participants dropped out and 70 were unreachable for the second interview. Additionally, 62 participants were excluded from the analysis because they contracted COVID-19 between the first interview and the completion of the second questionnaire. This resulted in a final sample of 639 participants included in the follow-up analysis. Among them, 438 tested positive for SARS-CoV-2. Further details are provided in Figure 1.

**Figure 1.**
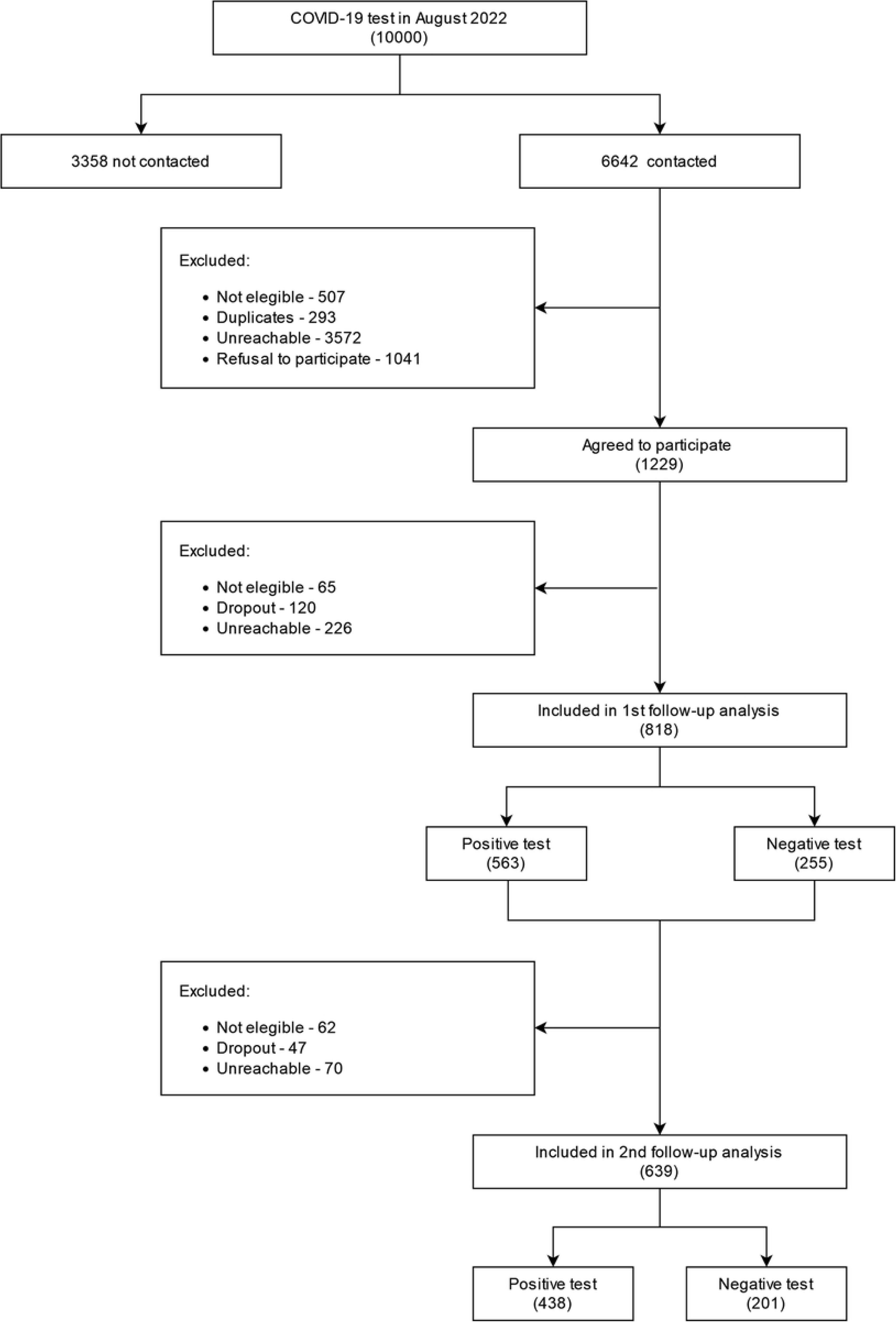
Flowchart of study participants.

A characterization of participants included at each time-point, considering sociodemographic, behavioral, and clinical characteristics prior to the SARS-CoV-2 test is present at table 1. At 9-month follow-up, those with positive tests tended to be younger (median 52 vs. 54 years old), female (58.3% vs. 53.7%), possessed higher education levels (46.4% vs. 35.7%), and were employed (67.1% vs. 54.9%). Participants who practiced regular physical exercise (≥30 minutes daily) were more common among negative cases (38.9% vs. 43.9%). Individuals testing positive tended to exhibit a higher frequency of alcohol consumption and the majority of participants across both groups reported consuming alcohol 2 to 4 times a month or less. Among the 639 participants who completed the 12- month follow-up, those with positive tests similarly tended to be younger (median 53 vs. 55 years old), more often female (58.5% vs. 50.8%), possessed higher education levels (46.6% vs. 37.8%), and were employed (65.30% vs. 58.7%). Consistent with findings from the first time-point, participants who practiced regular physical exercise were more common among negative cases (38.1% vs. 45.3%). Alcohol consumption exhibited an identical distribution pattern to the first follow-up. Regarding pre-existing medical conditions, prior COVID-19 infection and a history of psychiatric conditions were more common among individuals in the positive group at both 9 (21.9% vs. 15.7%; 17.2% vs. 13.7%, respectively) and 12 months (21.7% vs. 14.4%; 18.3% vs. 13.4%, respectively). On average, participants from both follow-ups with a positive SARS-CoV-2 test reported the presence of six symptoms at the time of the test, while participants with a negative test reported one symptom. The proportion of hospitalised participants, among those who tested positive, was residual at both time-points (≍ 1%). Moreover, the results regarding vaccination status were consistent across all groups and time-points, with each group having an average of three vaccines administered.

**Table 1.** Characterisation of the participants sociodemographic and clinical characteristics and vaccination status before the SARS-CoV-2 test.

| Variable | 9-months follow-up |  | 12-months follow-up |  |
| --- | --- | --- | --- | --- |
|  | N= 818 |  | N= 639 |  |
|  | COVID-19 test result |  |  |  |
|  | Negative, N(%) | Positive, N(%) | Negative, N(%) | Positive, N(%) |
|  | 255 (31.2%) | 563 (68.8%) | 201 (31.5%) | 438 (68.5%) |
| Age |  |  |  |  |
| Mean (SD) | 54 (18, 89) | 52 (18, 92) | 55 (18, 89) | 53 (18, 92) |
| Median (IQR) | 54 (43, 68) | 52 (41, 64) | 55 (45, 68) | 53 (42, 66) |
| Sex |  |  |  |  |
| Female | 137 (53.73%) | 328 (58.26%) | 102 (50.75%) | 256 (58.45%) |
| Male | 117 (45.88%) | 229 (40.67%) | 98 (48.76%) | 176 (40.18%) |
| Missing | 1 (0.39%) | 6 (1.07%) | 1 (0.50%) | 6 (1.37%) |
| Education level |  |  |  |  |
| Medium, primary education or lower education | 90 (35.29%) | 141 (25.04%) | 70 (34.83%) | 111 (25.34%) |
| Secondary education | 74 (29.02%) | 159 (28.24%) | 55 (27.36%) | 121 (27.63%) |
| Higher education | 91 (35.69%) | 261 (46.36%) | 76 (37.81%) | 204 (46.58%) |
| Missing | 0 (0.00%) | 2 (0.36%) | 0 (0.00%) | 2 (0.46%) |
| **Worker** |  |  |  |  |
| No | 115 (45.10%) | 184 (32.68%) | 82 (40.80%) | 151 (34.47%) |
| Yes | 140 (54.90%) | 378 (67.14%) | 118 (58.71%) | 286 (65.30%) |
| Missing | 0 (0.00%) | 1 (0.18%) | 1 (0.50%) | 1 (0.23%) |
| **Alcohol consumption** |  |  |  |  |
| Never | 93 (36.47%) | 157 (27.89%) | 66 (32.84%) | 127 (29.00%) |
| 2 to 4 times a month or less | 105 (41.18%) | 262 (46.54%) | 87 (43.28%) | 198 (45.21%) |
| 2 times a week or more | 57 (22.35%) | 144 (25.58%) | 48 (23.88%) | 113 (25.80%) |
| **Physical exercise (≥30 minutes daily)** |  |  |  |  |
| No | 143 (56.08%) | 343 (60.92%) | 110 (54.73%) | 270 (61.64%) |
| Yes | 112 (43.92%) | 219 (38.90%) | 91 (45.27%) | 167 (38.13%) |
| Missing | 0 (0.00%) | 1 (0.18%) | 0 (0.00%) | 1 (0.23%) |
| **Previous COVID-19 infection** |  |  |  |  |
| No | 214 (83.92%) | 431 (76.55%) | 172 (85.57%) | 334 (76.26%) |
| Yes | 40 (15.69%) | 123 (21.85%) | 29 (14.43%) | 95 (21.69%) |
| Unknown | 1 (0.39%) | 7 (1.24%) | 0 (0.00%) | 7 (1.60%) |
| Missing | 0 (0.00%) | 2 (0.36%) | 0 (0.00%) | 2 (0.46%) |
| **Previous psychiatric condition** |  |  |  |  |
| No | 220 (86.27%) | 463 (82.24%) | 174 (86.57%) | 355 (81.05%) |
| Yes | 35 (13.73%) | 97 (17.23%) | 27 (13.43%) | 80 (18.26%) |
| Missing | 0 (0.00%) | 3 (0.53%) | 0 (0.00%) | 3 (0.68%) |
| **N° of symptoms experienced in the 7 days before the SARS-CoV-2 test** |  |  |  |  |
| Mean (SD) | 1.33 (0.00, 12.00) | 5.61 (0.00, 16.00) | 1.27 (0.00, 12.00) | 5.66 (0.00, 16.00) |
| Median (IQR) | 0.00 (0.00, 1.00) | 5.00 (3.00, 8.00) | 0.00 (0.00, 1.00) | 5.50 (3.00, 8.00) |
| **Level of care needed** |  |  |  |  |
| Hospitalised | NA | 3 (0.53%) | NA | 2 (0.46%) |
| Non-hospitalised | NA | 554 (98.40%) | NA | 431 (98.40%) |
| Missing | NA | 6 (1.07%) | NA | 5 (1.14%) |
| **N° of COVID-19 vaccine doses** |  |  |  |  |
| Mean (SD) | 3.00 (0.00, 5.00) | 2.99 (0.00, 5.00) | 3.07 (0.00, 5.00) | 3.01 (0.00, 5.00) |
| Median (IQR) | 3.00 (3.00, 4.00) | 3.00 (2.75, 4.00) | 3.00 (3.00, 4.00) | 3.00 (3.00, 4.00) |
| Missing | 1 (0.39%) | 3 (0.53%) | 1 (0.50%) | 3 (0.68%) |

Notes: NA – Non-applicable.

Our study analyzed the prevalence of each outcome among individuals tested for SARS-CoV-2, as detailed in Figure 2 and Supplementary Table 1, and complemented this with a difference of proportion analysis (Table 2). Memory loss showed a higher prevalence among individuals who tested positive (prevalence [Pr]: 24.87, 95% confidence interval [CI]: 21.35; 28.65) compared to those who tested negative (Pr: 10.20, 95%CI: 6.77; 14.58) at the 9-month follow-up, with a significant difference of proportion between prevalences (14.81, 95% CI: 9.35; 20.27). Similarly, among participants in the subsequent survey, memory loss was more prevalent in those with a positive test result (Pr: 16.67, 95% CI: 13.30; 20.49) compared to those with a negative result (Pr: 9.45, 95% CI: 5.79; 14.37), with a difference of proportions of 7.33 (95% CI: 1.61; 13.05). Concentration issues were also more prevalent among participants with a positive test result, both at the 9-month follow-up (Pr: 15.45, 95% CI: 12.57; 18.71 vs. Pr: 7.45, 95% CI: 4.55; 11.39), with a difference of proportions of 8.08 (95% CI: 3.39; 12.79), and at the 12-month follow-up (Pr: 9.82, 95%CI: 7.20; 13.00 vs. Pr: 2.99, 95%CI: 1.10; 6.38), with a difference of proportions of 6.83 (95% CI: 2.82; 10.84). Examining the prevalence of both symptoms, the presence of at least one cognitive symptom at 9 months was more prevalent in the positive test group (Pr: 28.24, 95% CI: 24.56; 32.16) compared to the negative test group (Pr: 12.16, 95% CI: 8.41; 16.81), with a difference of proportions of 16.14 (95% CI: 10.36; 21.91). At the 12-month follow-up, the prevalence was still higher in the positive test group (Pr: 17.81, 95% CI: 14.34; 21.72) compared to the negative test group (Pr: 9.95, 95% CI: 6.18; 14.95), with a difference of proportions of 7.90 (95% CI: 2.06; 13.74). Moreover, the prevalence of having both symptoms was higher in the positive group at 9 months (Pr: 12.08, 95% CI: 9.50; 15.06 vs. Pr: 5.49, 95% CI: 3.03; 9.04), with a difference of proportions of 6.61 (95% CI: 2.43; 10.79), and at the 12-month follow-up (Pr: 8.68, 95% CI: 6.21; 11.71 vs. Pr: 2.49, 95% CI: 0.81; 5.71), with a difference of proportions of 6.19 (95% CI: 2.42; 9.95).

**Figure 2.**
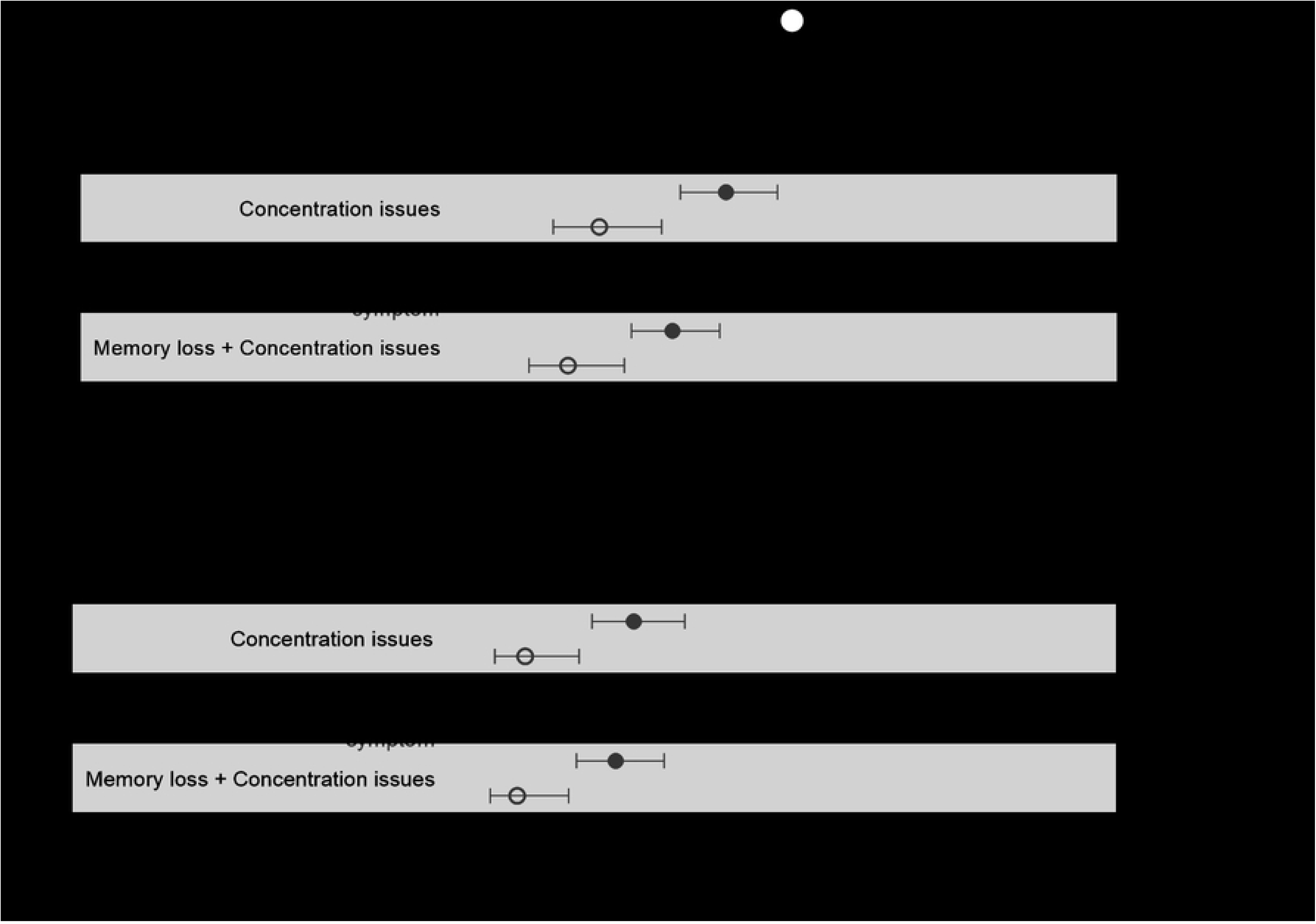
Prevalence of cognitive symptoms 9 and 12 months after SARS-CoV-2 test by test result.

**Table 2.**
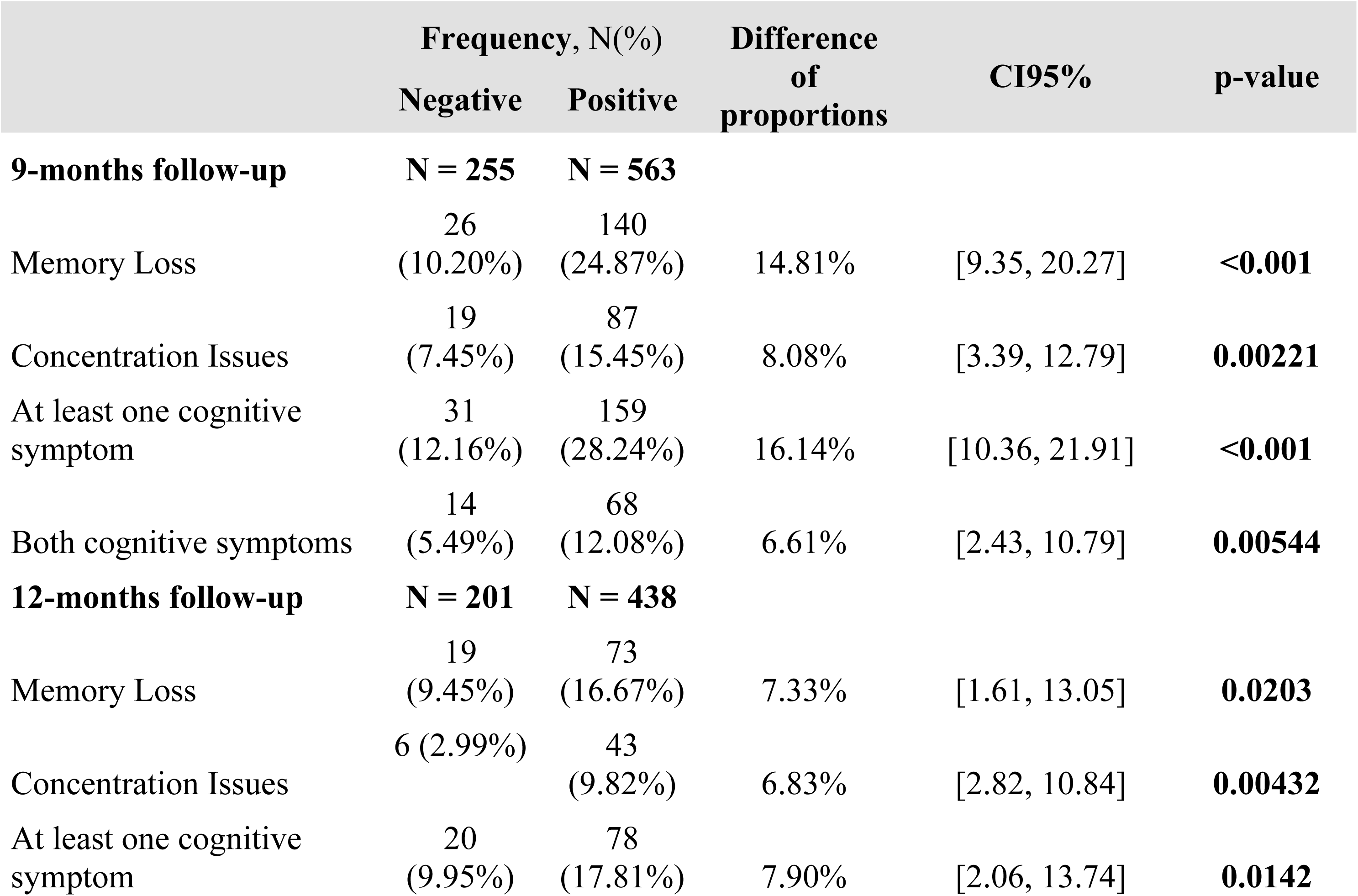
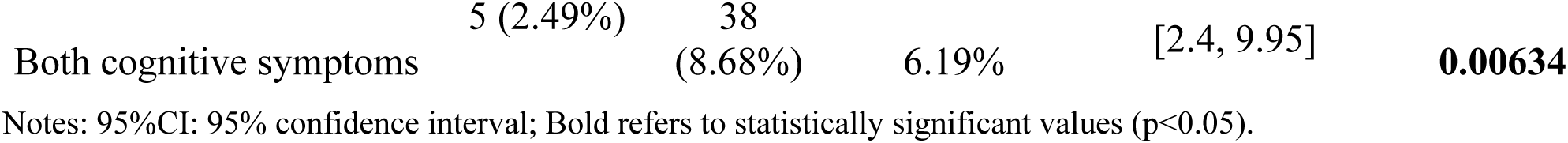
Difference of proportions analysis for cognitive symptoms at 9-month and 12-month follow-ups.

|  | Frequency, N(%) |  | Difference of proportions | CI95% | p-value |
| --- | --- | --- | --- | --- | --- |
|  | Negative | Positive |  |  |  |
| **9-months follow-up** | **N = 255** | **N = 563** |  |  |  |
| Memory Loss | 26<br>(10.20%) | 140<br>(24.87%) | 14.81% | [9.35, 20.27] | **<0.001** |
| Concentration Issues | 19<br>(7.45%) | 87<br>(15.45%) | 8.08% | [3.39, 12.79] | **0.00221** |
| At least one cognitive symptom | 31<br>(12.16%) | 159<br>(28.24%) | 16.14% | [10.36, 21.91] | **<0.001** |
| Both cognitive symptoms | 14<br>(5.49%) | 68<br>(12.08%) | 6.61% | [2.43, 10.79] | **0.00544** |
| **12-months follow-up** | **N = 201** | **N = 438** |  |  |  |
| Memory Loss | 19<br>(9.45%) | 73<br>(16.67%) | 7.33% | [1.61, 13.05] | **0.0203** |
| Concentration Issues | 6 (2.99%) | 43<br>(9.82%) | 6.83% | [2.82, 10.84] | **0.00432** |
| At least one cognitive symptom | 20<br>(9.95%) | 78<br>(17.81%) | 7.90% | [2.06, 13.74] | **0.0142** |

Notes: 95%CI: 95% confidence interval; Bold refers to statistically significant values (p<0.05).

Furthermore, to assess the potential impact of age-related cognitive decline on our findings, we conducted a sensitivity analysis, excluding participants aged 60 years or older (see Supplementary Table 2 for complete analysis). We found similar trends regarding each symptom individually. Notably, the prevalence of having at least one cognitive symptom revealed a significantly higher prevalence in those with a positive test at 12-month follow-up (Pr: 11.87; 95%CI: 9.00; 15.28 vs. Pr: 4.48; 95%CI: 2.07; 8.33), supported by a difference of proportions of 11.33 (95% CI: 4.18; 18.49). Regarding the presence of both symptoms, a higher prevalence was still observed among the positive group at both the 9-month (Pr: 7.28; 95% CI: 5.28; 9.75 vs. Pr: 2.75; 95% CI: 1.11; 5.57) and 12-month (Pr: 5.48; 95% CI: 3.54; 8.04 vs. Pr: 1.00; 95% CI: 0.12; 3.55) follow-ups. When estimating the difference in proportions, we found that the disparity between the groups remained statistically significant for all studied outcomes at both time-points (Supplementary Table 3).

The comparison of cognitive symptoms between age groups (under 60; 60 and over) at both time-points is further depicted in Figure 3. Individuals aged 60 and over exhibit a higher prevalence of all outcomes except concentration issues and at least one cognitive symptom at the 12-month follow-up. Nevertheless, the overall distribution remains highly consistent across both age groups, with both groups showing a decline in the prevalence of all outcomes over time.

**Figure 3.**
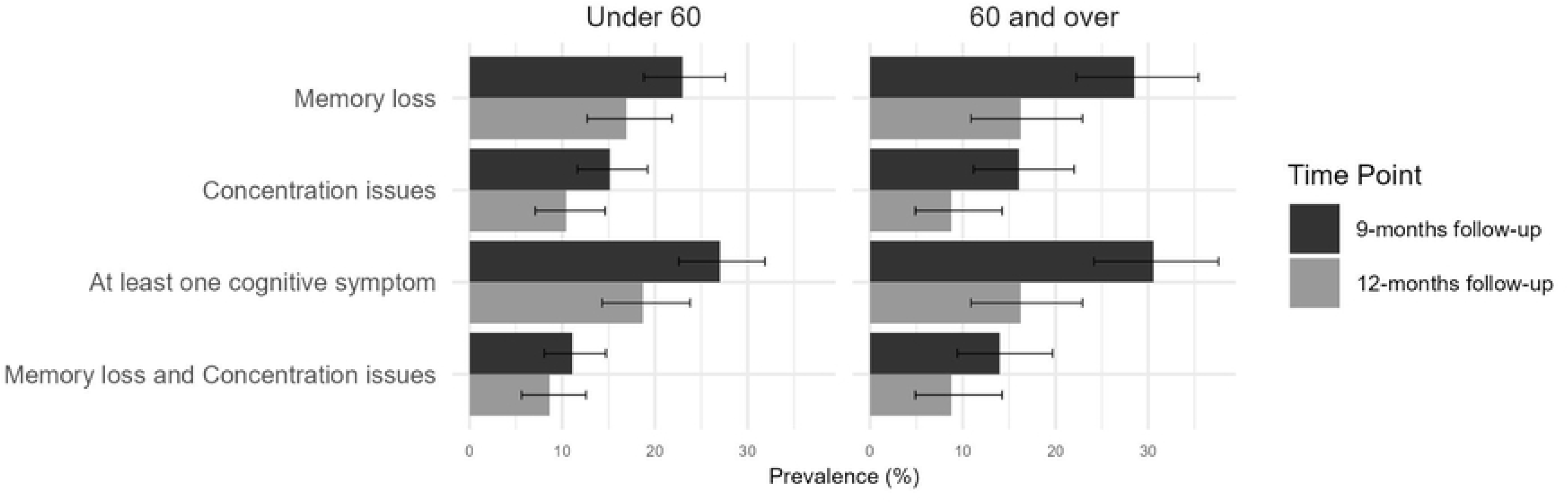
Prevalence of cognitive symptoms among positive cases 9 and 12 months after SARS-CoV-2 test by age group

We further analyzed the sociodemographic, behavioral, and clinical factors associated with having at least one cognitive symptom in participants who tested positive for SARS-CoV-2 (Table 3). Our adjusted results reveal a significant association between being female and post-COVID-19 cognitive symptoms, evident both at 9 months (Adjusted Prevalence Ratio [aPR]: 2.02, 95% CI: 1.42; 2.87) and 12 months (aPR: 1.97, 95% CI: 1.19; 3.26). The distribution of key demographic and health variables, including age, COVID-19 test results, education level, and previous COVID-19 infection status, between males and females at both the 9- and 12-month follow-ups is provided in Supplementary Table 4. Additionally, while a history of psychiatric condition showed significance in unadjusted results at 9 months (aPR: 1.50, 95% CI: 1.11; 2.02), this association did not maintain significance following adjustment. Analysis of the remaining variables (age, education level, alcohol intake, physical exercise, prior COVID-19 infection, and COVID-19 vaccine doses) revealed no significant association with post-COVID-19 cognitive symptoms.

**Table 3.** Factors associated with the prevalence of post-COVID cognitive symptoms.

| Variables | 9-months follow-up (N = 546) |  |  |  | 12-months follow-up (N = 422) |  |  |  |
| --- | --- | --- | --- | --- | --- | --- | --- | --- |
|  | Crude |  | Adjusted |  | Crude |  | Adjusted |  |
|  | PR | 95%CI | PR | 95%CI | PR | 95%CI | PR | 95%CI |
| **Sex** |  |  |  |  |  |  |  |  |
| Male | Ref. |  |  |  |  |  |  |  |
| Female | **1.95** | **[1.42, 2.67]** | **2.02** | **[1.42, 2.87]** | **1.62** | **[1.03, 2.54]** | **1.97** | **[1.19, 3.26]** |
| **Age** |  |  |  |  |  |  |  |  |
| Under 60 | Ref. |  |  |  |  |  |  |  |
| 60 and over | 1.1 | [0.83, 1.45] | 1.23 | [0.89, 1.17] | 0.8 | [0.51, 1.24] | 0.95 | [0.58, 1.57] |
| **Education level** |  |  |  |  |  |  |  |  |
| Higher education | Ref. |  |  |  |  |  |  |  |
| Medium, primary education or lower education | 0.94 | [0.67, 1.33] | 0.85 | [0.57, 1.26] | 0.61 | [0.34, 1.09] | 0.59 | [0.31, 1.12] |
| Secondary education | 1.04 | [0.76, 1.43] | 1.01 | [0.73, 1.38] | 0.96 | [0.61, 1.53] | 0.94 | [0.58, 1.53] |
| **Alcohol intake** |  |  |  |  |  |  |  |  |
| Never | Ref. |  |  |  |  |  |  |  |
| 2 to 4 times a month or less | 1.13 | [0.81, 1.57] | 1.21 | [0.87, 1.69] | 0.86 | [0.52, 1.43] | 0.87 | [0.51, 1.45] |
| Twice a week or more | 1.05 | [0.71, 1.53] | 1.38 | [0.92, 2.06] | 1.24 | [0.74, 2.10] | 1.62 | [0.90, 2.91] |
| **Physical exercise** |  |  |  |  |  |  |  |  |
| Yes | Ref. |  |  |  |  |  |  |  |
| No | 1.24 | [0.93, 1.65] | 1.18 | [0.88, 1.58] | 0.98 | [0.64, 1.49] | 0.94 | [0.61, 1.47] |
| **Previous COVID-19 infection** |  |  |  |  |  |  |  |  |
| No | Ref. |  |  |  |  |  |  |  |
| Yes | 1.14 | [0.84, 1.56] | 1.03 | [0.74, 1.43] | 1.28 | [0.81, 2.02] | 1.18 | [0.72, 1.93] |
| **Psychiatric condition** |  |  |  |  |  |  |  |  |
| No | Ref. |  |  |  |  |  |  |  |
| Yes | **1.50** | **[1.11, 2.02]** | 1.23 | [0.90, 1.68] | 1.08 | [0.65, 1.81] | 0.84 | [0.49, 1.44] |
| **COVID-19 vaccine doses** | 0.99 | [0.86, 1.15] | 0.97 | [0.82, 1.14] | 0.85 | [0.70, 1.03] | 0.85 | [0.68, 1.07] |

Notes: PR: Prevalence ratio; 95%CI: 95% confidence interval; physical exercise was defined as $\geq 30$ minutes daily; Bold refers to statistically significant values.

## 4 Discussion

One of the key findings of this study is the higher prevalence of memory loss and concentration issues among individuals who tested positive for SARS-CoV-2 compared to those who tested negative at both 9-month and 12-month follow-up. Furthermore, in participants who tested positive, we found that being female was associated with a higher prevalence of Long COVID symptoms at both time-points.

Our results show that at 9 months memory loss and concentration issues had a prevalence rate of 24.9% and 15.5%, respectively. At 12 months, prevalence rates decreased but were still more prevalent among positive cases, with 16.7% of the participants reporting memory loss and 9.8% reporting concentration issues. This finding aligns with previous research reporting a prevalence between 11.0% and 34.5% for memory loss and 2.6% and 31.0% for concentration issues(Ahmed et al., 2022; Fernández-de-las-Peñas et al., 2023; Garrigues et al., 2020; Keijsers et al., 2022; Merza et al., 2023; Pilotto et al., 2021; Søraas et al., 2021). Differences in study designs, follow-up periods, collection procedures, and populations, may explain the heterogeneous prevalence rates among studies. For instance, studies with hospitalized patients often show higher prevalences of cognitive post-COVID complications, as these symptoms seem to manifest more in patients with initially severe disease(Fernández-de-Las-Peñas et al., 2022; Garrigues et al., 2020; Keijsers et al., 2022; Pérez-González et al., 2022). Fernández-de-las-Peñas et al.(Fernández-de-Las-Peñas et al., 2022) showed that two years after SARS-CoV-2 infection, memory loss was reported by 20.0% of hospitalized patients versus 15.9% of non-hospitalized patients. However, these symptoms are not exclusive to severe cases and have also been noted in patients with initially mild COVID-19 infection, emphasizing the importance of studies focused on mild cases, such as ours, where less than 1.0% of participants required hospitalization. Pilotto et al.(Pilotto et al., 2021) showed that among mild cases 33.3% reported memory and/or concentration problems at 6-months follow-up. Similarly, in our study, among those testing positive, the prevalence of having at least one cognitive symptom was 28.2% at the 9-month follow-up, dropping to 17.8% at 12 months, with a difference of proportions of 16.14 and 7.90, respectively. Other studies also showed a tendency for cognitive symptoms to decline over time following infection[17,18]. Baeler et al.[18] implemented an online quiz to investigate working memory following COVID-19 infection. Their study revealed a gradual increase in memory scores over a 17-month period post-COVID-19, indicating a decrease in cognitive impairment over time. Another study indicated that out of the 14.9% of patients reporting memory loss approximately 8 months post-infection, only 5.8% continued to experience these symptoms at 12 months[17]. However, the same study revealed that approximately 6.0% of participants who did not exhibit memory loss at the initial assessment developed it 12 months after infection. This highlights the importance of investigating post-COVID cognitive symptoms, especially beyond the one-year mark following infection.

Moreover, most studies evaluating post-COVID cognitive symptoms overlook the impact of age-related cognitive decline. This cognitive decline may start as early as in one’s 20s and 30s(Salthouse, 2009). However, the pace of cognitive decline tends to accelerate, especially after the age of 60(Hedden & Gabrieli, 2004). Despite evidence suggesting older individuals are more prone to memory impairment as a post-COVID symptom, this remains debated among researchers(Baseler et al., 2022; Merza et al., 2023). The sensitivity analysis excluding participants aged 60 years or older revealed that among younger individuals with a positive SARS-CoV-2 test, our results showed similar trends, with memory loss being more prevalent at the 9-month follow-up and concentration issues more prevalent at both time points compared with those who tested negative for SARS-CoV-2. This analysis demonstrated a reduced influence of age-related cognitive decline on our findings, highlighting the consistency of our results. Moreover, although participants aged 60 or over showed a slightly higher prevalence of all analyzed outcomes compared to their younger counterparts, the overall distribution across age groups was very similar, with both groups showing a notable decline in all outcomes over time. These findings indicate that while older adults may be more susceptible to certain cognitive impairments, younger individuals were also impacted by these post-COVID complications. Future research should continue to explore these age-related differences better to tailor post-COVID care and support for diverse age groups.

Additionally, we analyzed individual factors associated with post-covid cognitive symptoms in participants with a positive test result. Being female emerged as a significant risk factor for developing post-covid cognitive symptoms, consistent with previous studies(Merza et al., 2023) highlighting gender disparities in COVID-19 outcomes. While an interaction analysis between cognitive symptoms and sex should be further explored, we believe that studies with larger cohorts would be better suited to investigate this potential interaction. Additionally, older age, previous COVID-19 infection and not being vaccinated have also been reported as risk factors(Merza et al., 2023; Pilotto et al., 2021) for post-covid symptoms, however, no significant association was found in our results. Several factors may contribute to this discrepancy, including variations in sample characteristics, such as the lack of hospitalized participants and those with severe disease typically present in studies identifying these risk factors(Al-Aly et al., 2021; Merza et al., 2023; Pilotto et al., 2021), as well as methodological differences in study design, such as the reliance on self-reported symptoms which may limit the accuracy of the reported symptom prevalence. Additionally, potential limitations in statistical power and the influence of unaccounted confounding variables, such as comorbidities, may also affect the results. Moreover, vaccination, the severity of acute infection, and symptom presentation (symptomatic vs asymptomatic) may represent crucial risk factors identified in the literature(Al-Aly et al., 2021; Jennings et al., 2023; Merza et al., 2023; Pilotto et al., 2021) that were not fully addressed in our study due to data limitations, specifically the low number of participants with severe cases, the small number of participants without vaccination, and insufficient representation of asymptomatic individuals. These aspects underscore the need for further research with larger, more diverse cohorts to better understand the complex interplay between risk factors and the development of post-COVID cognitive symptoms.

Furthermore, comorbidities might influence the development of post-COVID symptoms, yet specific conditions associated with post-COVID memory loss and concentration issues remain poorly understood. Multiple studies have demonstrated that pre-existing mental health conditions can affect the severity of acute disease and elevate the risk of developing post-COVID symptoms(Greißel et al., 2024; Hüfner et al., 2022; Wang et al., 2021). Therefore, we hypothesize that a prior history of psychiatric conditions may increase the risk of post-COVID cognitive symptoms. While our findings indicate an association between previous psychiatric conditions and post-COVID cognitive symptoms at the 9-month follow-up, this relationship lost significance after adjusting for other variables. However, it is worth noting that the proportion of participants with a history of these conditions may be underestimated, as mental health disorders are often underdiagnosed due to stigma, limited access to mental health services, and challenges in recognizing and accurately diagnosing symptoms, which can be misunderstood, overlooked, or miscommunicated in clinical settings(Clement et al., 2015; Corrigan et al., 2014; Kasper, 2006; Milton & Mullan, 2014). Thus, further research is essential to explore the impact of pre-existing mental health conditions on post-COVID cognitive impairment. It is crucial to acknowledge that our study possesses certain limitations that deserve careful attention. A significant limitation of our study pertains to the control group. Although we used a control group consisting of individuals with negative test results, reliance on negative test results does not guarantee the absence of prior infections. At both time-points, around 15% and 21% of participants with negative and positive SARS-CoV-2 tests, respectively, reported having a prior COVID-19 diagnosis. Furthermore, we were only able to include a lower number of negative participants. This could influence the outcomes and reduce the robustness of our findings. Moreover, the relatively small sample size, especially notable during the second follow-up, may have failed to demonstrate some associations between sociodemographic and clinical characteristics and post-COVID cognitive symptoms. Additionally, given that our sample predominantly consists of adults and older adults, we may have overlooked the prevalence distribution across other younger age groups. Furthermore, the small number of hospitalized individuals in our sample poses a limitation on the incorporation of this factor into our regression analysis. This is particularly significant as numerous studies have consistently highlighted acute disease severity as a key risk factor contributing to the development of cognitive symptoms subsequent to COVID-19 infection(Al-Aly et al., 2021; Merza et al., 2023; Pilotto et al., 2021). However, even though we may lose some diversity regarding specific variables like hospitalization or severity status, the use of a community-based sample in our study offers significant advantages. Unlike clinical samples, which may be biased toward individuals seeking treatment, community-based samples include individuals regardless of their health-seeking behavior, providing a more representative and comprehensive picture of the population.

Considering the methodology used, most studies investigating post-COVID cognitive symptoms use specific cognitive and memory assessment tools and tests that target various memory and cognitive functions, complicating direct comparisons across studies. Furthermore, different terminologies, such as “brain fog” and “cognitive impairment,” encompass a range of symptoms that vary across studies, leading to divergent interpretations, particularly with self-reported symptoms. In our study, we applied a WHO-approved questionnaire (ISARIC) and participants were queried about symptoms experienced in the preceding 7 days, which were absent before their SARS-CoV-2 test. However, it is crucial to recognize that reported symptoms might also relate to conditions other than COVID-19, such as flu-like symptoms that could emerge following the test, whether diagnosed or undiagnosed. Symptoms were self-reported, relying on participants’ comprehension, recollection, understanding of symptom definitions, and personal assessment. To enhance clarity, we used plain language and avoided medical terminology. Another constraint was our inability to thoroughly analyze the vaccination variable while considering the vaccination dates, as we originally intended. This was due to data limitations and a significant amount of missing information regarding the timing of COVID-19 vaccination (whether administered post or prior infection), which is known to be a factor associated with Long COVID symptoms, including cognitive symptoms(Byambasuren et al., 2023; Jennings et al., 2023). Still considering the methodology used, we assessed symptoms at 9 and 12 months post-SARS-CoV-2 test. These intervals are particularly noteworthy in light of emerging research suggesting that cognitive symptoms might only surface as late as one year after infection(Fernández-de-las-Peñas et al., 2023).

Our study significantly enhances the understanding of post-COVID cognitive symptoms in Portugal and addresses a crucial gap by comparing the prevalence of memory loss and concentration issues, both individually and in their co-presence, among individuals who tested positive and negative for SARS-CoV-2. Additionally, we tested SARS-CoV-2-negative individuals in the same month as the positive group, minimizing time disparities between the two cohorts, unlike other studies that might use different infections or individuals with no history of COVID-19 infection as the control group. Furthermore, many studies investigating cognitive symptoms neglect the impact of age-related cognitive decline. Our sensitivity analysis bridged this gap, allowing us to examine the prevalence of post-COVID cognitive symptoms in a younger sample. This enhanced the robustness of our results and revealed that younger people are also significantly affected by these post-COVID complications. As such, their inclusion in future research and incorporation into public health strategies is crucial.

In summary, our analysis aimed to assess the prevalence of memory loss and concentration issues between participants who tested positive and negative for SARS-CoV-2 and the factors associated with post-COVID cognitive symptoms within SARS-CoV-2 positive test participants. Despite challenges due to sample size, our results offer valuable insights. In a predominantly non-hospitalised population, we found a higher prevalence of cognitive symptoms in patients with positive SARS-CoV-2 test, peaking at 9 months post-infection and declining thereafter. Moreover, we show that these symptoms, often overlooked and attributed to age-related conditions, are not exclusive to older populations, underscoring the need for broader consideration. This evidence is crucial for clinicians, who should be aware that these symptoms can occur in individuals with non-severe COVID-19 and may pose a significant burden in everyday life.

## Data Availability

Data cannot be shared publicly due to the confidentiality of participant information. However, researchers who meet the criteria for access to confidential data can request access by signing a declaration to carry out research projects using the LOCUS database. Requests should be directed to the project email.

## 5 Conflict of Interest

The authors declare that the research was conducted in the absence of any commercial or financial relationships that could be construed as a potential conflict of interest.

## 6 Author Contributions

CR, MM, PS, AL contributed to the design of this study. CR, MM, and AL contributed to data collection. Data preparation and statistical analysis were performed by CR with oversight from MM, AL and PS. The first draft of the manuscript was produced by CR with feedback from all other authors. All authors reviewed, edited, and approved the final version.

## 7 Funding

This study was sponsored by Pfizer (grant code #68639655; URL: https://www.pfizer.pt/). The funders did not have a role in study design, data collection and analysis, decision to publish or preparation of the manuscript.

## 8 Acknowledgments

The authors thank Direção-Geral da Saúde and Serviços Partilhados Ministério da Saúde for data sharing and Pfizer for funding. We also thank all the participants for their valuable time and to the interviewers for their perseverance, which was essential to collect these data.

## 9 Ethics approval

This study involved human participants and was approved by the Ethics Committee for Health of the Regional Health Administration of Lisbon and Tagus Valley (2151/CES/2022) and the Data Protection Officer of the General Directorate of Health. Verbal informed consent was obtained from the participants prior to the questionnaire.

## 12 LOCUS group

André Peralta Santos, Andreia Costa, Andreia Vilas-Boas, António Carlos da Silva, Gabriel Atanásio, Inês Simões, Joana Paixão, João V. Cordeiro, João Victor Rocha, Lelita Santos, Luísa Eça Guimarães, Maria da Luz Brazão, Maria João Lobão, Mário Santos, Marta Sofia Fonseca, Patrícia Barbosa, Sofia Nóbrega, Sónia Dias and Víctor Ramos.

## Notes

### Competing Interest Statement

The authors have declared no competing interest.

### Clinical Protocols

https://journals.plos.org/plosone/article?id=10.1371/journal.pone.0285051

